# Low-Dose Whole-Lung Radiation for COVID-19 Pneumonia

**DOI:** 10.1101/2020.07.11.20147793

**Authors:** Clayton B. Hess, Zachary S. Buchwald, William Stokes, Tahseen H. Nasti, Jeffrey M. Switchenko, Brent D. Weinberg, Nadine Rouphael, James P. Steinberg, Karen D. Godette, David Murphy, Rafi Ahmed, Walter J. Curran, Mohammad K. Khan

**Affiliations:** Departments/Divisions of Radiation Oncology, Emory University. Atlanta GA; Departments/Divisions of Microbiology and Immunology, Emory University. Atlanta GA; Departments/Divisions of Biostatistics and Bioinformatics, Emory University. Atlanta GA; Departments/Divisions of Infectious Disease, Emory University. Atlanta GA; Departments/Divisions of Radiology, Emory University. Atlanta GA; Departments/Divisions of Pulmonary, Allergy, Critical Care and Sleep Medicine, Emory University. Atlanta GA; Departments/Divisions of Winship Cancer Institute. Emory University. Atlanta GA

## Abstract

**Background:** Safety of whole-lung low-dose radiation therapy (LD-RT) for COVID-19 pneumonia has been established in two phase I trials. By focally dampening pulmonary cytokine hyperactivation, LD-RT may improve outcomes in hospitalized and oxygen-dependent COVID-19 patients.

**Methods:** Patients with COVID-19 pneumonia were treated with 1.5 Gy whole-lung LD-RT, followed for 28 days or at least until hospital discharge, and compared to an age- and comorbidity-matched control cohort. COVID-19-positive patients eligible for this protocol were hospitalized, had radiographic consolidations, and required supplemental oxygen. Efficacy endpoints were time to clinical recovery, radiographic improvement, and serologic responses.

**Results:** Ten patients received whole-lung LD-RT between April 24 and May 24, 2020 and were compared to ten matched control patients, of whom six received COVID-directed therapy. Median time to clinical recovery was 12 days for the control cohort vs 3 days for LD-RT (HR 2.9, p=0.05). Median time to hospital discharge (20 and 12 days, p=0.19), and intubation rates (40% and 10%, p=0.12) were shorter for the LD-RT cohort. The LD-RT cohort had faster radiographic improvement (p=0.03), even among patients with high COVID burden. Serologic recovery in specific hematologic, cardiac, hepatic, clotting, and inflammatory markers occurred more rapidly following LD-RT than among matched controls.

**Conclusions:** Strong efficacy signals, including a 3-fold risk reduction in time to clinical improvement, were observed following LD-RT compared to matched patients receiving COVID-directed therapy for COVID-19 pneumonia. Given the global availability of radiation accelerators, ongoing international efforts to investigate the optimal role of LD-RT in COVID-19 pneumonia are justified.

**Clinical Trial Registration:** NCT04366791.

## Introduction

The novel coronavirus discovered in 2019 (COVID-19) has brought unprecedented global death and disruption. While most infected patients exhibit an indolent course, those with advanced age or comorbidities face higher risk of respiratory failure, mediated by a cascading hyperinflammatory macrophage activation event in the lungs^2^, and face a mortality of 30-80% once dependent on mechanical ventillation.^3-5^ SARS-CoV-2 viral particles infect alveolar type II pneumocytes and produce a cascading inflammatory event in the airway.^6^ This cytokine storm can lead to pulmonary edema, infiltrative inflammatory cells, diffuse alveolar damage, and injury to myocardium and extra-pulmonary organs.^7-9^

Anti-Inflammatory effects of low-dose radiation therapy (LD-RT) can result in apoptosis and decrease adhesion of the leukocytes to endothelial cells,^10^ mitigate proinflammatory effects of macrophages by reducing secretion of nitric oxide and reactivation oxygen species,^11^ and induce polarization of M1-inflammatory macrophages to M2-anti-inflammatory subtype.^12^ Furthermore, LD-RT may also reduce TNF-α, IL-1β, IL-2, IL-6, IL-8, and INF-γ,^13-17^ which are thought to be increased in the COVID-19 cytokine storm. Thus, it was hypothesized that LD-RT directed at the lungs may dampen the cytokine hyperactivation and improve the outcome of COVID-19 hospitalized and oxygen-dependent patients. Seven-day interim phase I safety data from the first five patients treated on this trial was reported and established LD-RT as a safe intervention with no detected acute toxicity or exacerbation of the cytokine storm.^1^ The full efficacy data from the first known prospective trial of LD-RT for COVID-19 pneumonia compared with a matched control cohort is presented.

## Methods

### Trial Design

The Radiation Eliminates Storming Cytokines and Unchecked Edema as a 1-day Treatment for COVID-19 (RESCUE 1-19) trial is an investigator-initiated, single-institution combined phase I/II trial aimed to determine safety and then to explore preliminary efficacy of single-fraction, whole-lung LD-RT for hospitalized and oxygen-dependent patients with COVID-19 pneumonia. Clinical Trial Registration Number NCT04366791. The research protocol was approved by the Emory University Institutional Review Board. All participants gave written informed consent prior to any study procedures. The study protocol and approved addenda permitted treatment of an initial cohort of five pre-intubated patients with a planned 7-day interim analysis and safety stopping rule to evaluate acute toxicity and cytokine storm exacerbation. After evaluating safety, an institutional data safety monitoring committee permitted investigators to proceed with five additional treatments to evaluate efficacy. A total of 10 pre-intubated patients received LD-RT and were followed for a minimum of 28 days or until discharge. A cohort of age- and comorbidity-matched controls was selected from COVID-positive patients previously enrolled on another prospective institutional trial for comparative outcome analysis. Study investigators were blinded to the selection and outcomes of control patients, who were matched by age and comorbidity burden.

### Patients

Eligible LD-RT patients were positive for COVID-19 by nasopharyngeal swab using polymerase chase reaction (PCR)-based testing, were hospitalized, had pneumonic consolidation on either x-ray or computed tomographic (CT) imaging, required oxygen supplementation, and were assessed by providers as clinically declining (altered mental status, increasing oxygen demands, and/or weaning intolerance). Exclusion criteria included actual or planned pregnancy or administration of COVID-directed drug therapies within one day prior to radiotherapy delivery through post-LD-RT day 3. Anti-pyretic medications were suspended at enrollment. Following LD-RT, clinical staff was instructed to attempt oxygen weaning as clinically indicated in non-declining patients at no less than 12-hour intervals, while maintaining oxygen saturations above 90%. Patients were pre-planned for clinical assessment at the time of enrollment and on post-RT days 1, 3, and 7, and 28, as well as optional assessment on days 14 and 21. The Glasgow Coma Scale (GCS)^18^ and Charlson Comorbidity Index (CCI)^19^ were used to assess mental status and comorbidity burden, respectively. Radiographs were permitted at any time as clinically indicated but obtained per-protocol at least 12 hours prior to radiation, 24 hours following radiation, and on post-RT days 3, 7, 28, and optionally at day 14 and 21. Evaluation of serum inflammatory, renal, cardiac, chemistry, clotting, and hematologic markers were encouraged daily, but obtained at least at baseline and also on post-RT days 3, 7, and 28, and optionally on days 14 and 21. Age as a binary variable was added to the analysis plan to evaluate time to clinical recovery in patients age 65 and older compared to patients age 64 and younger based on observations made during the trial.

Ten control patients from another IRB-approved institutional protocol were selected and matched for age and comorbidity for comparison with the LD-RT cohort, for evidence of efficacy. Eligible control patients also tested COVID positive and were selected from enrollees on a prospective institutional phlebotomy protocol for outbreaks of diseases of public health importance, including COVID-19. Controls were permitted but not required to be co-enrolled on any trial of COVID-19-directed therapies, including the Adaptive COVID-19 Treatment Trial (ACTT-1, Clinical Trial NCT04280705).

### Intervention

Enrolled patients received best supportive care plus LD-RT to a dose of 1.5 Gy to the bilateral whole lungs, delivered in a single fraction, utilizing a 2-dimensional therapeutic radiation technique, an anterior-posterior beam configuration, and standard dose rates. Patients in the control cohort received best supportive care with or without COVID-directed therapies (ie, remdesevir, hydroxychloroquine, glucocorticosteroids, etc.) per protocol or physician discretion. Time of COVID-directed therapy start was defined as the date of LD-RT delivery (in the radiation cohort), as the first day of administration of COVID-directed therapies (in control patients if received), or as the first full-day of hospitalization (in control patients who received best supportive care alone).

### Outcome Measures

The trial’s primary objective of safety was reported previously for the first five patients who received LD-RT.^1^ The endpoint of efficacy is measured by time to clinical recovery, defined as the time from LD-RT administration to the first day on which a subject satisfies one of three categories from an ordinal scale: (1) Not hospitalized, no limitations on activities; (2) Not hospitalized, limitation on activities and/or requiring home oxygen; or (3) Hospitalized, not requiring supplemental oxygen. Additional secondary outcomes related to clinical course, radiographic improvement, and serology. Clinical course was evaluated as time from intervention to hospital discharge, total hospital duration, intubation events, duration of intubation, oxygenation requirements, and vital status. Radiographic responses were measured by serial imaging. Chest x-rays were evaluated as improved (I), stable (S), or worse (W) by a board-certified diagnostic radiologist (BW) compared to baseline and also blindly assigned an ordinal 1-5 score, using a severe acute respiratory syndrome (SARS) scoring scale without knowledge of cohort designation or timing of intervention.^20^ Chest computed tomography obtained at baseline and day 7 were subjectively assessed for 3-dimensional radiographic amongst LD-RT patients. Serological efficacy was measured by serial laboratory evaluations.

#### Statistical Analysis

Two-sample t-tests and chi-squared tests were used for continuous and categorical endpoints, respectively. Cumulative incidence of recovery and discharge were plotted using the Kaplan-Meier method. Patients not recovered or intubated were censored at date of death or hospital discharge. Univariate Cox proportional hazards models were fit, and hazard ratios were reported. Serial imaging SARS scores were carried forward from Day 7 to 14 to 21 if missing. Median and interquartile range was calculated for laboratory values at clustered time points: 3 days before RT through the day of intervention, and at days 1-3, 4-7, and 8-14 post-RT, when available.

## Results

### Patients

From April 23 to May 24, 2020, fifty-one patients were screened for eligibility, and thirteen were enrolled to the LD-RT cohort. Three patients became ineligible for transport due to COVID symptom worsening and clinical decline prior to radiation delivery (1 died, and 2 were intubated). The remaining ten were treated with LD-RT (Figure 1, supplemental). Ten control patients, who had been admitted between March 27 and May 12, 2020 and enrolled on another institutional prospective trial, were matched for a comparative analysis based on age and comorbidities. Table 1 outlines patient demographic at the time of hospital admission. Median age was 78 (range 43-104) and 75 (44-99) for the LD-RT and control cohorts, respectively (p=0.06). Seventy-five percent were African-American, 55% were female, and 40% were residents of nursing homes that experienced COVID-19 infection outbreaks. Median CCI comorbidity scores were 6.5 (range 0-10) and 5.0 (0-8), respectively (p=0.19). Median duration of symptoms prior to admission was 7.5 (range 1-30) and 5.5 (0-21) days, respectively (p=0.33). One control patient was admitted for COVID positivity but was asymptomatic on presentation. Median GCS scores were 14 (range 8-15) and 15 (14-15) on admission, and mild (range 13-15) in 70% and 100% of each cohort, respectively (p=0.16). Median oxygen supplementation requirement at the time of admission were 3 liters (range 0-15) and 2 liters (0-15), respectively (p=0.46). Common presenting symptoms on admission for the whole cohort were dyspnea/cough (65%), fever/chills (45%), dizziness/confusion/altered mentation (40%), and body aches/myalgias/weakness (25%). Patients received LD-RT later in their hospital stay (median day 4.5, range 1-16) than controls received COVID-directed intervention (median day 2.0, range 1-4, p=0.06). Patients age 65 and over had less severe oxygen dependence (median 3 liters/min) at the time of COVID-directed intervention compared to younger patients (6 liters/min, p=0.05). Median documented follow up was 22 days in the LD-RT cohort compared to 46 days in the control cohort.

**Table 1.**
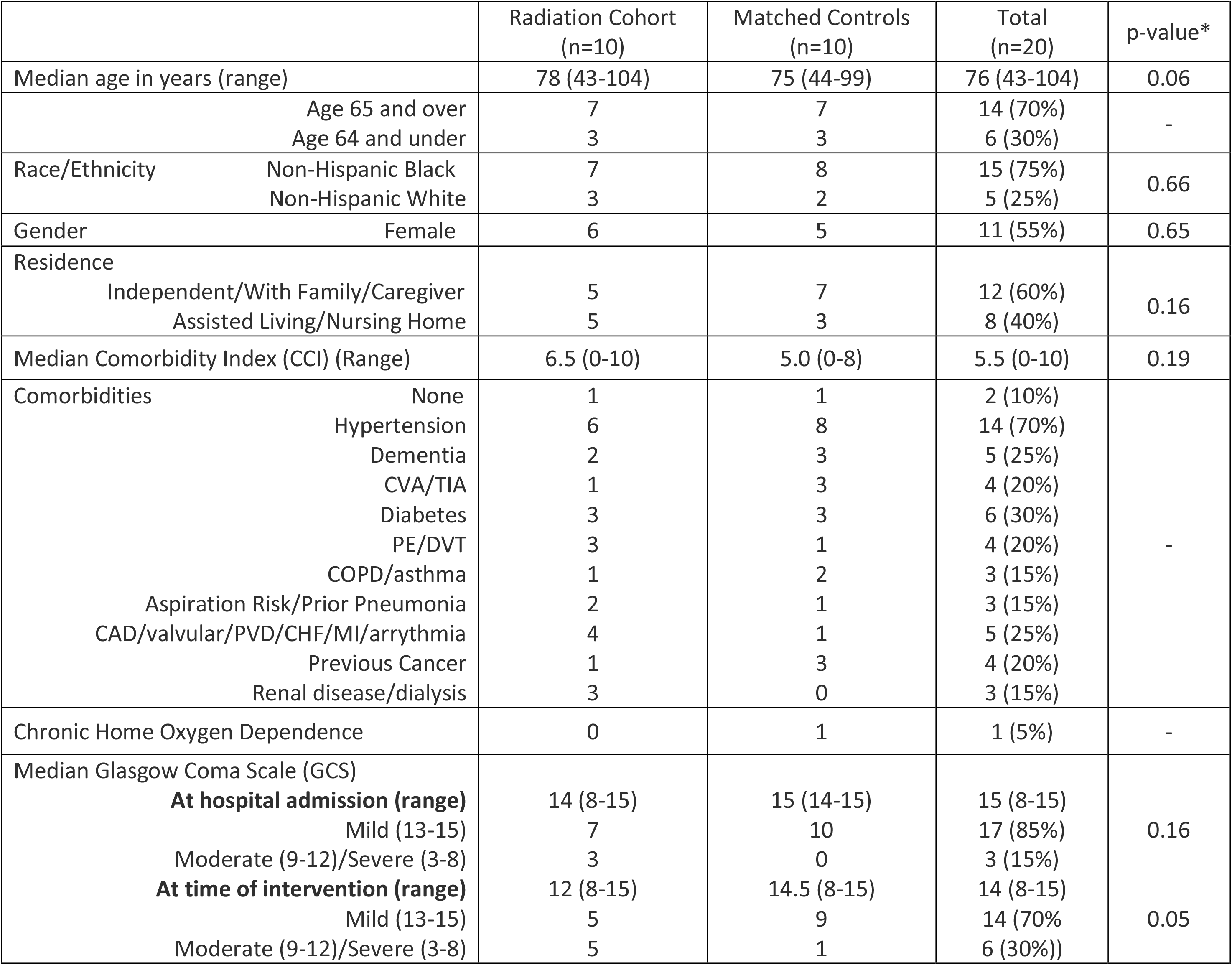
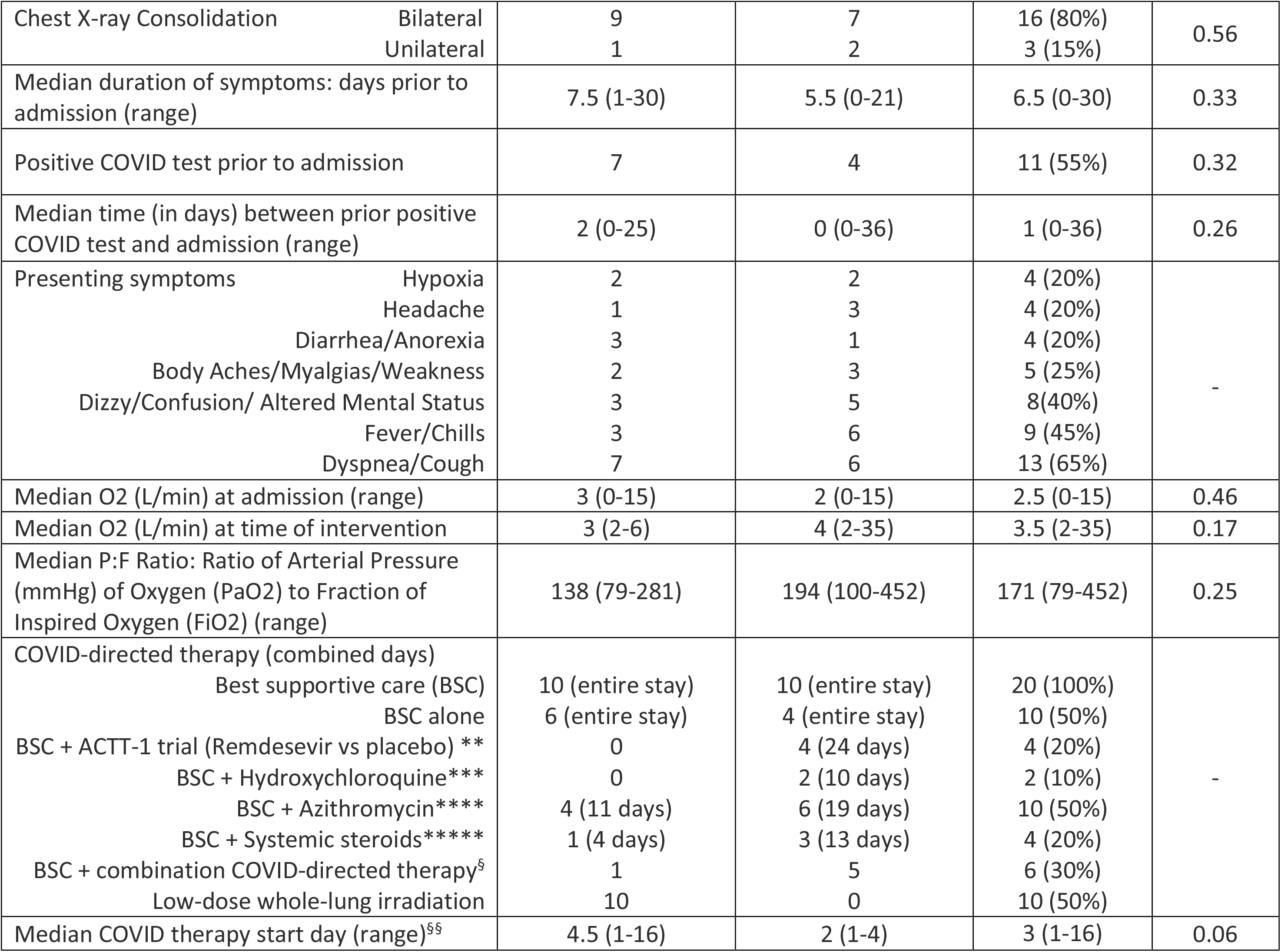
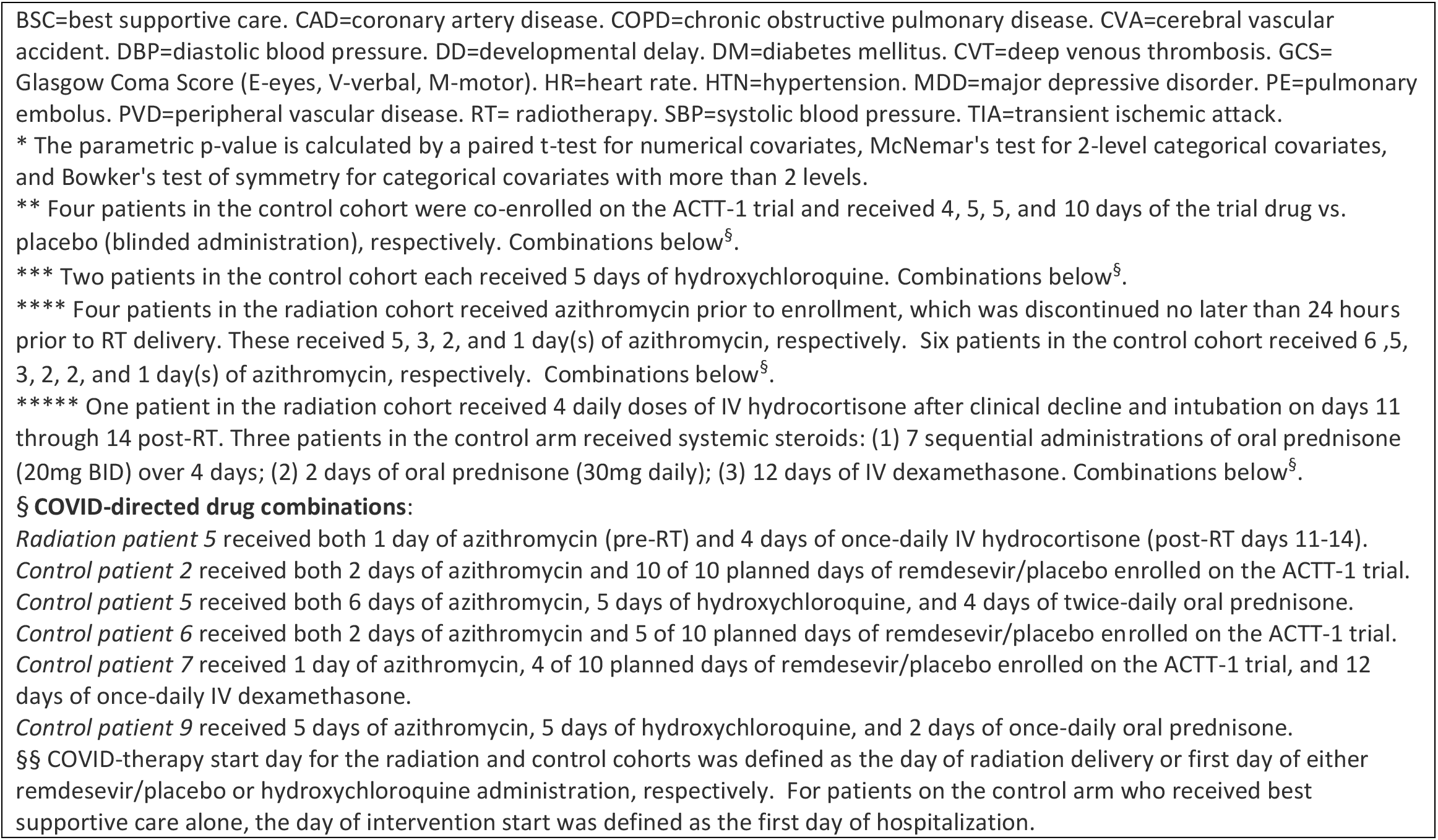
Patient Demographics

**Figure 1.**
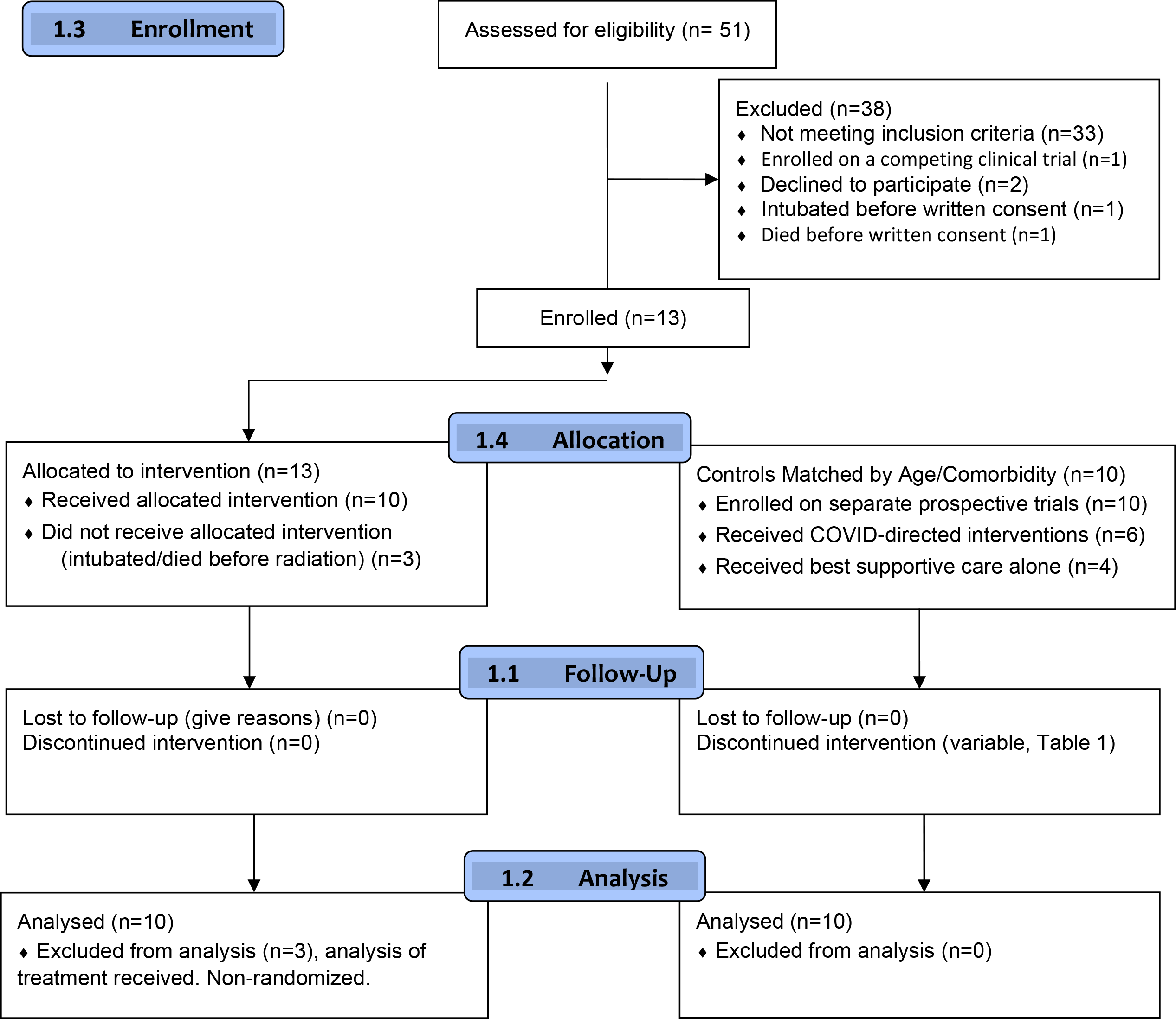
CONSORT Flow Diagram (Supplemental)

### Clinical Outcomes

Median time to clinical improvement was 3 days (range 3 hours to 8.5 days) in the LD-RT cohort compared to 12 days (range 19 hours to 32 days) in the control cohort (HR 2.9, CI 1.0-8.39, p=0.05, Figure 2a). Median time from COVID-directed therapy to hospital discharge was 12 days (range 7 to 25) compared to 20 days (5 to 45 days), respectively (HR 2.13, CI 0.68-6.66, p=0.19, Figure 2b). Freedom from intubation was 90% and 60%, respectively (p=0.11, Figure 2c). Additional treatment outcomes are reported in Table 2. Median time from admission to hospital discharge was 16 days (range 13 to 42) compared to 19 days (7 to 45), respectively (p=0.56). Twenty-eight-day overall survival was 90%, and median survival time was not reached in both cohorts. Median days intubated was 4.3 and 1 day(s), respectively (p=0.12). Median total time requiring oxygen supplementation was 10 days (range 4-18) in LD-RT cohort compared to 13 days (range 1-33) in the control cohort (p=0.15). Age 65 and over was associated with a lower oxygen requirement at the time of intervention and was associated with a shorter time to clinical recovery in the LD-RT cohort (p=0.01) but not the control cohort (p=0.40).

**Table 2.**
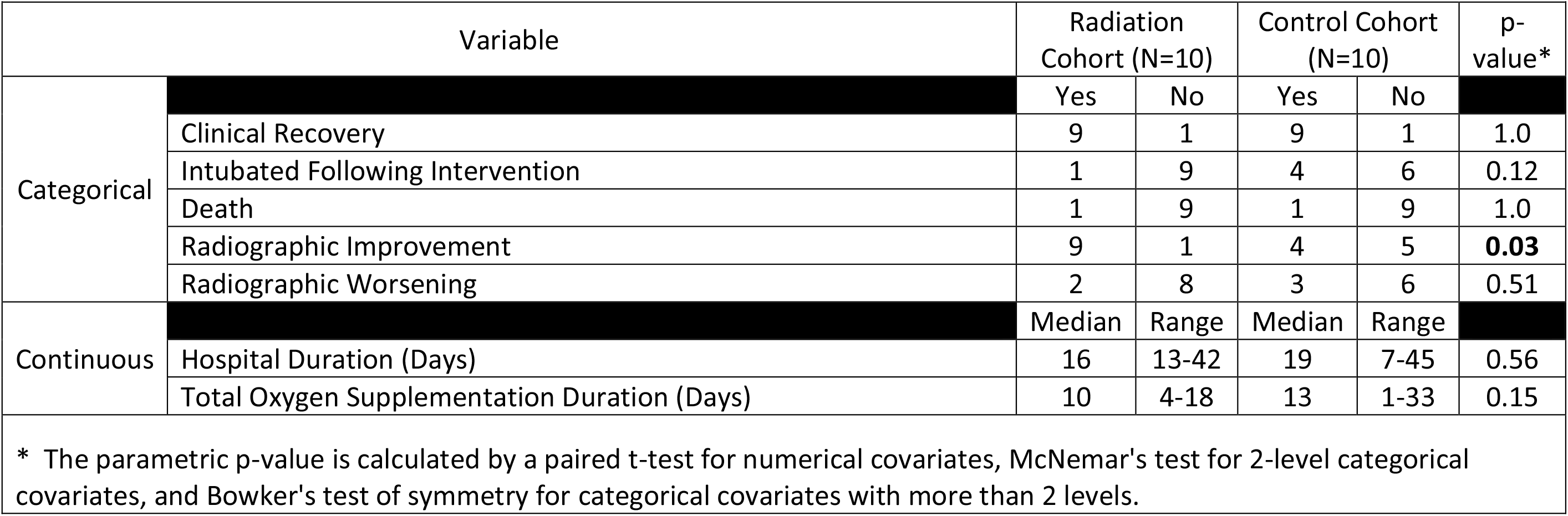
Treatment Outcomes

**Figure 2a.**
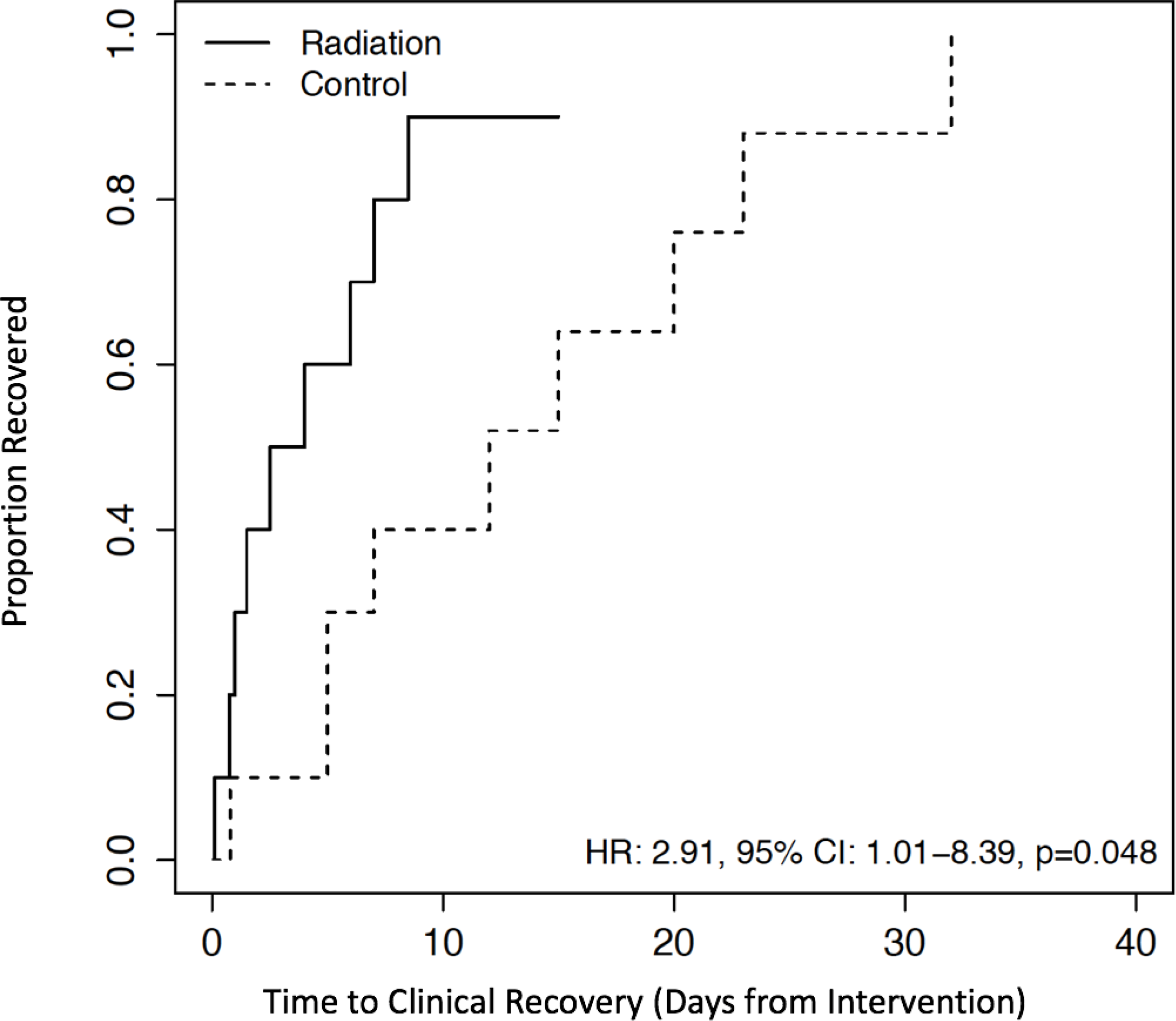
Time from COVID-19 Directed Therapy to Clinical Recovery

**Figure 2b.**
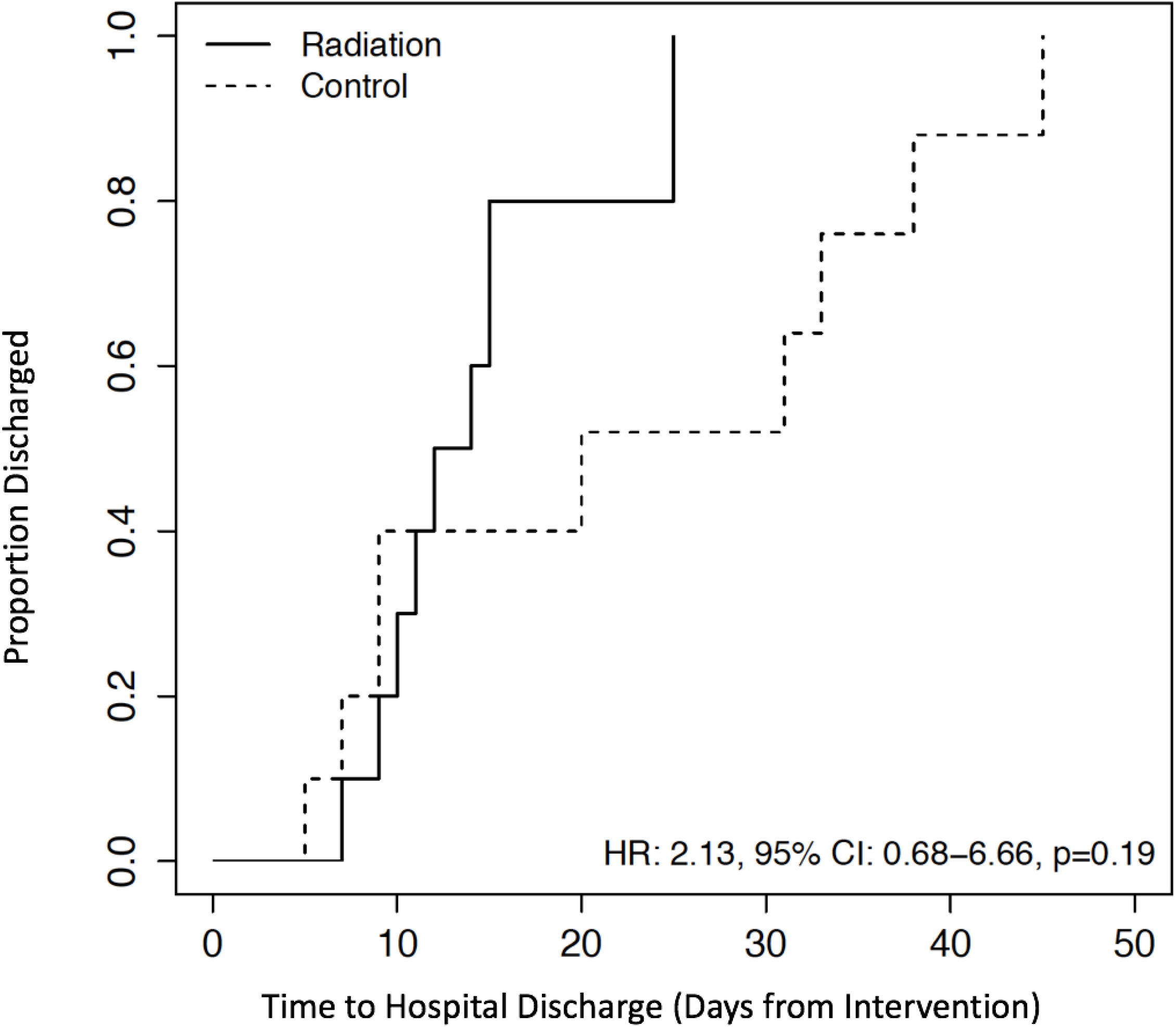
Time From COVID-19 Directed Therapy Start to Hospital Discharge

**Figure 2c.**
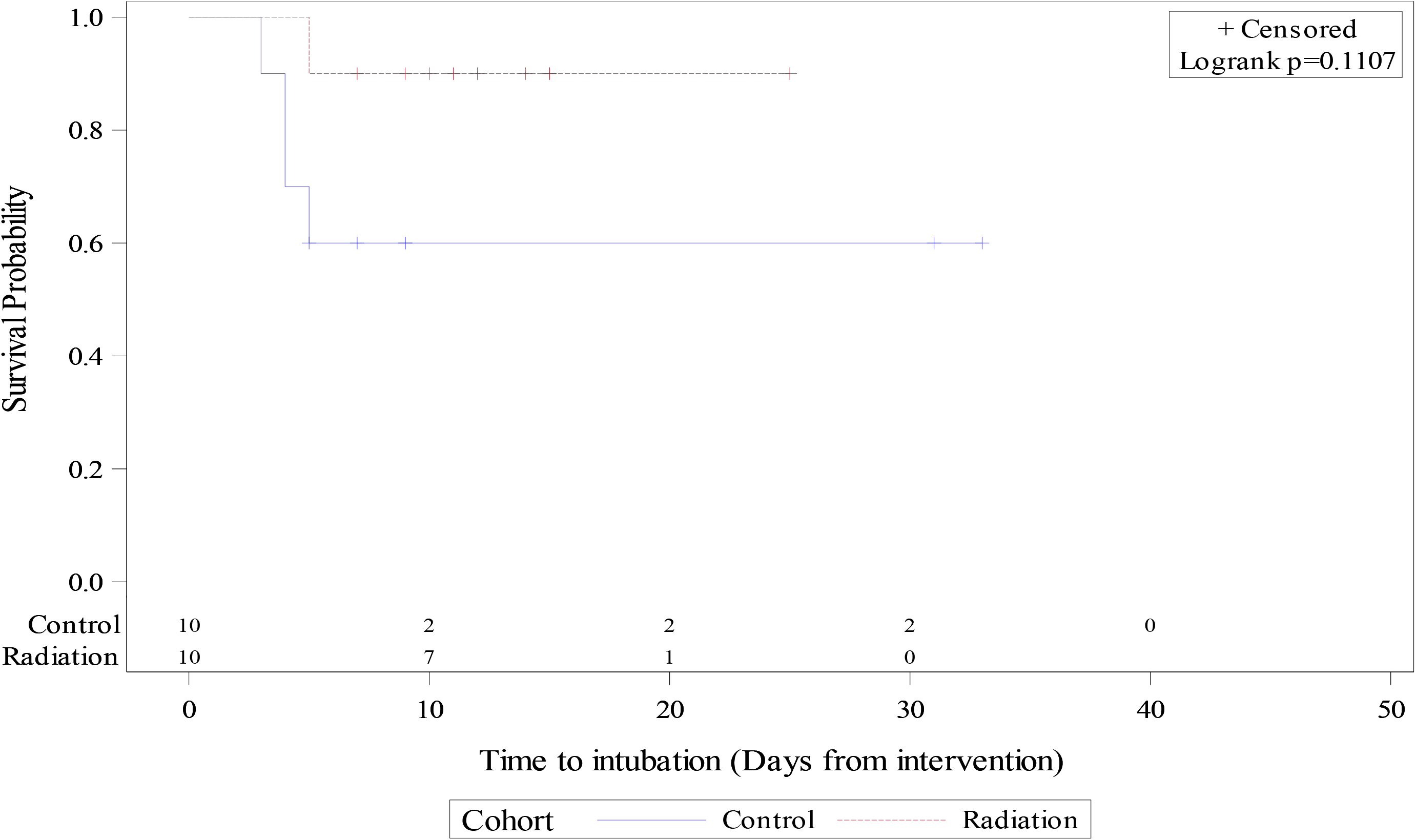
Time to Intubation

### Mentation

Patients in the LD-RT cohort had more severe mental status changes (median GCS 12 vs. 14.5, p=0.33) and more commonly a GCS score of 10 or lower (50% compared to 10%, p=0.05) at the time of COVID-directed intervention. Within 24 hours of COVID-directed therapy, change in median GCS was 2.5 points higher (range 0 to 5) in the LD-RT cohort compared to controls (p<0.01), whose GCS was stable in all but one patient who cognitively declined.

### Radiographic Response

Radiographic improvement was more common in the LD-RT cohort (p=0.03, Table 2). Clinical vignettes of patients with high burden of pulmonary COVID and corresponding 3-dimensional radiographic responses to LD-RT on computed tomography are shown in Figure 3. Average daily ARDS scale scores for serial x-rays from all patients are shown in Figure 4. Mean change in radiographic ARDS scale between baseline and last available x-ray at day 21 was superior in the LD-RT cohort compared to controls (p=0.17, Figure 4).

**Figure 3.**
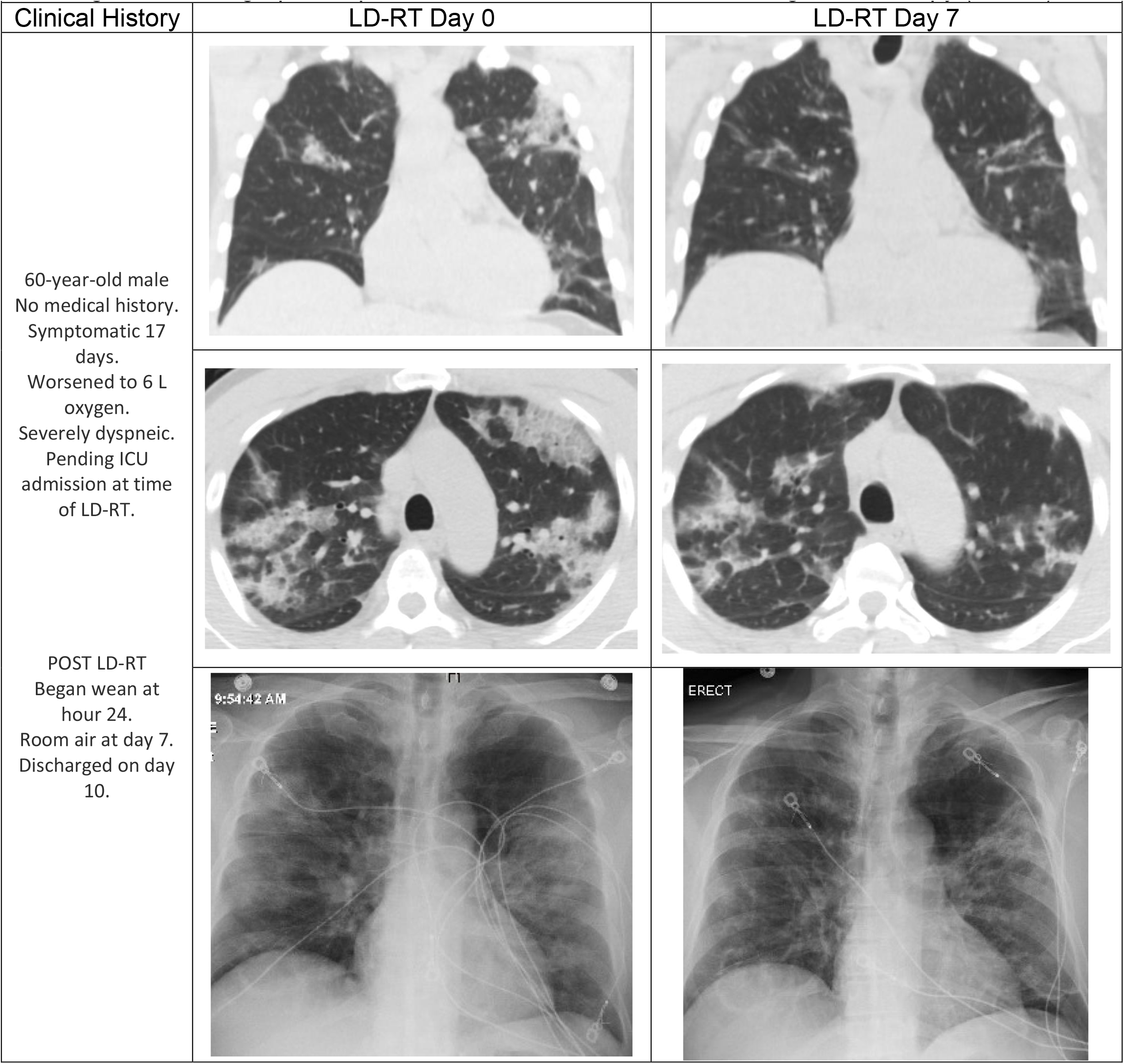
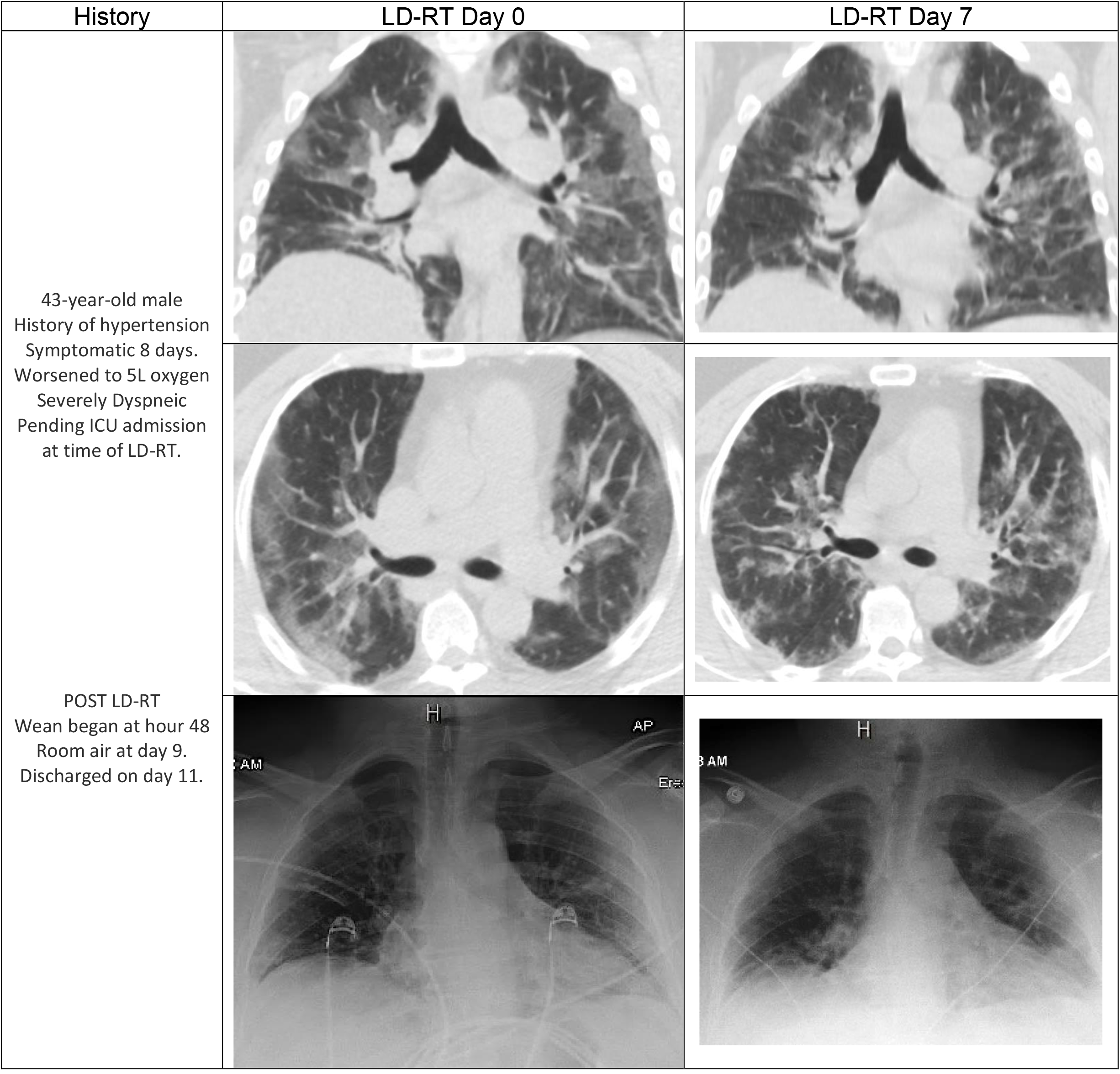
Radiographic Improvement After Low-Dose Whole-Lung Radiotherapy (LD-RT)

**Figure 4.**
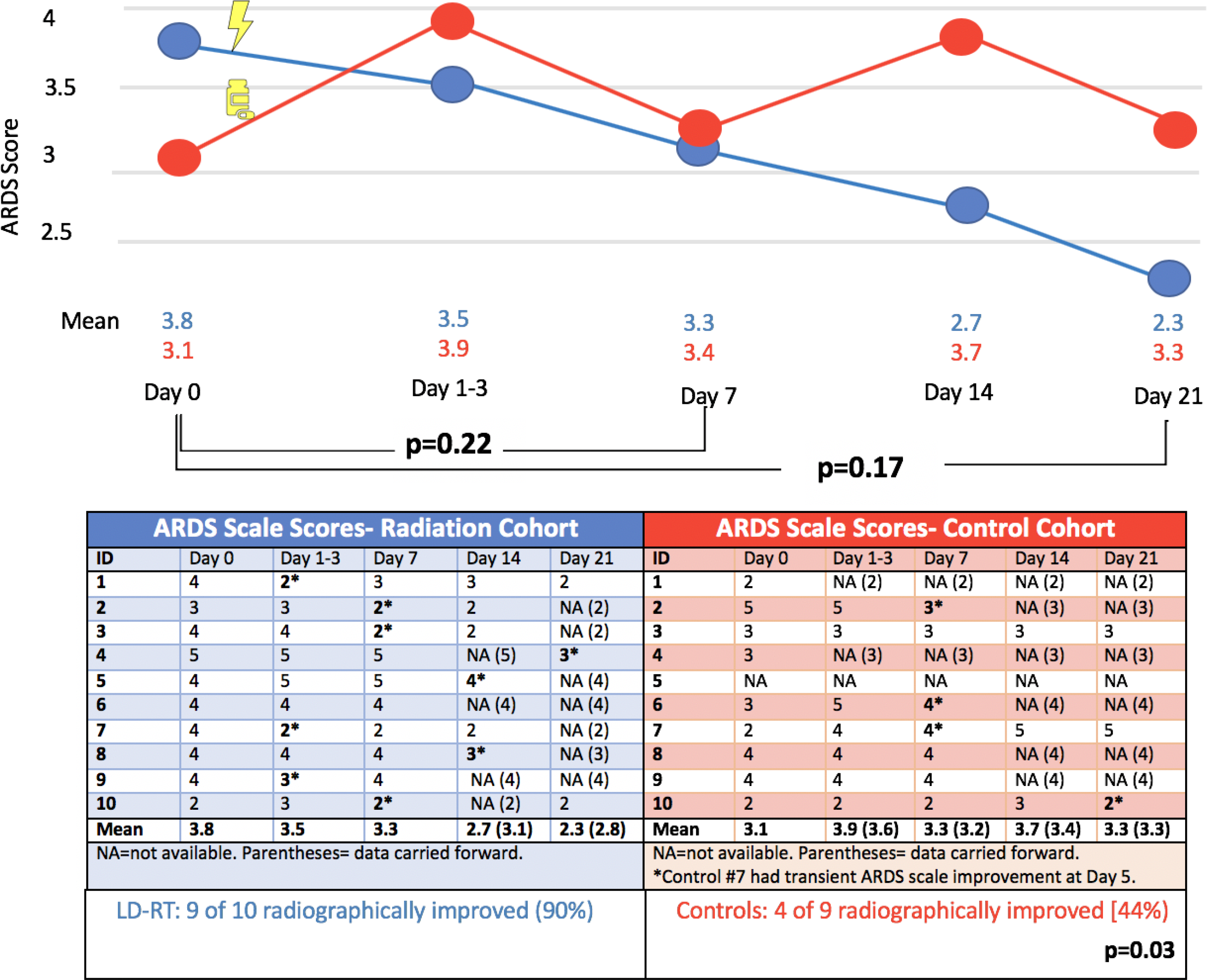
ARDS X-ray scale scores pre- and post-COVID directed intervention

### Serologic Response

Safety of hematologic, renal, cardiac, chemistry, clotting, and inflammatory markers within 7 days following LD-RT was reported previously.^1^ Comparison of medians and inter-quartile ranges of LD-RT patients to controls is shown in Figure 5 (supplemental). Improvement over time and/or statistical difference between baseline and post-LD-RT day 7 was observed for c-reactive protein (p=0.17), lactate dehydrogenase (p=0.04), creatine kinase (p=0.94), d-dimer (p=0.27), troponin (p=0.17), AST (p=0.12), ALT (p=0.06), and white blood cells count (p<0.01). Creatinine levels did not differ over time between the cohorts (p=0.80). Interleukin-6, myoglobin, fibrinogen, erythrocyte sedimentation rate, ferritin, and procalcitonin also trended downward, and control levels were not available in controls for comparison.

**Figure 5.**
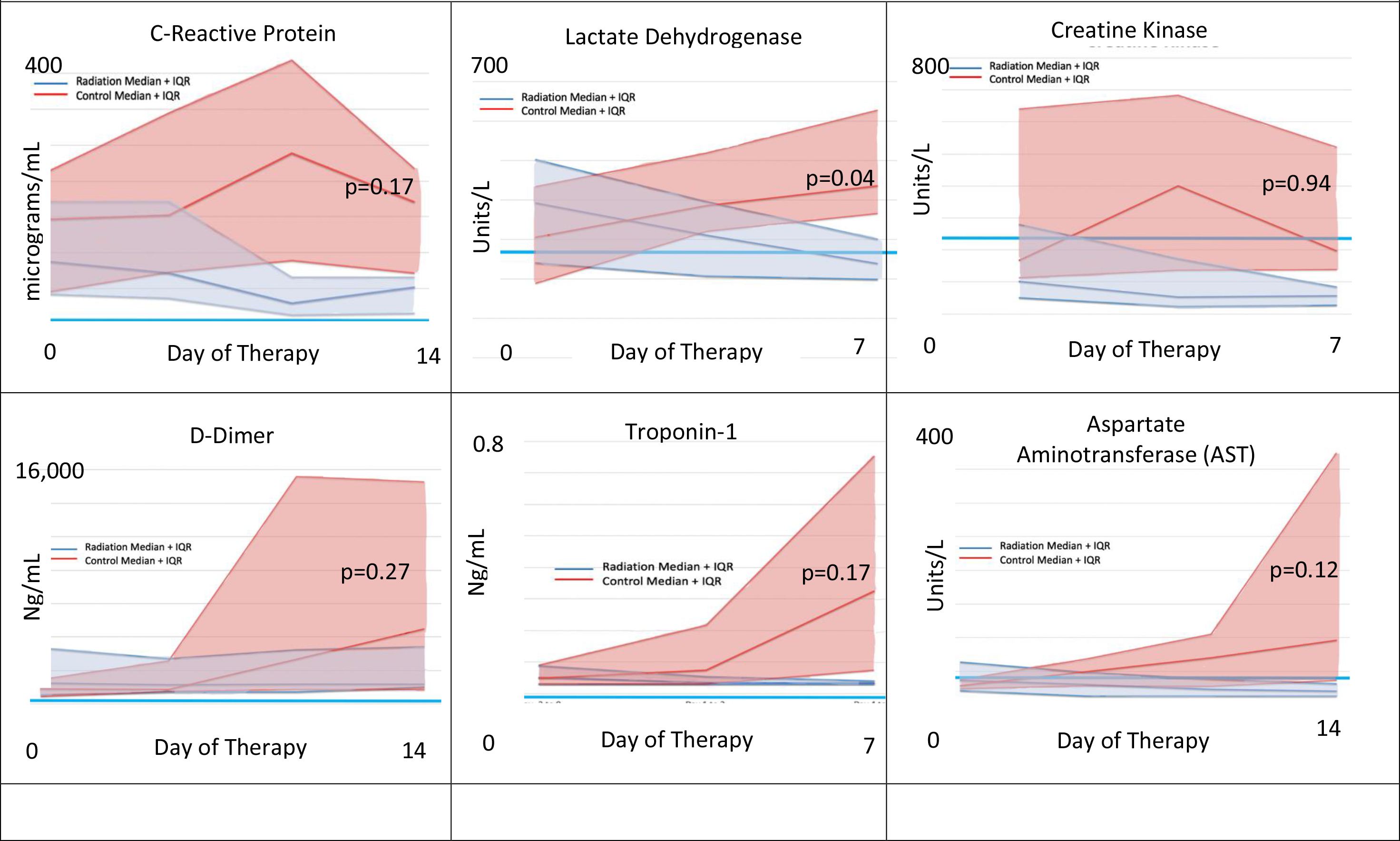
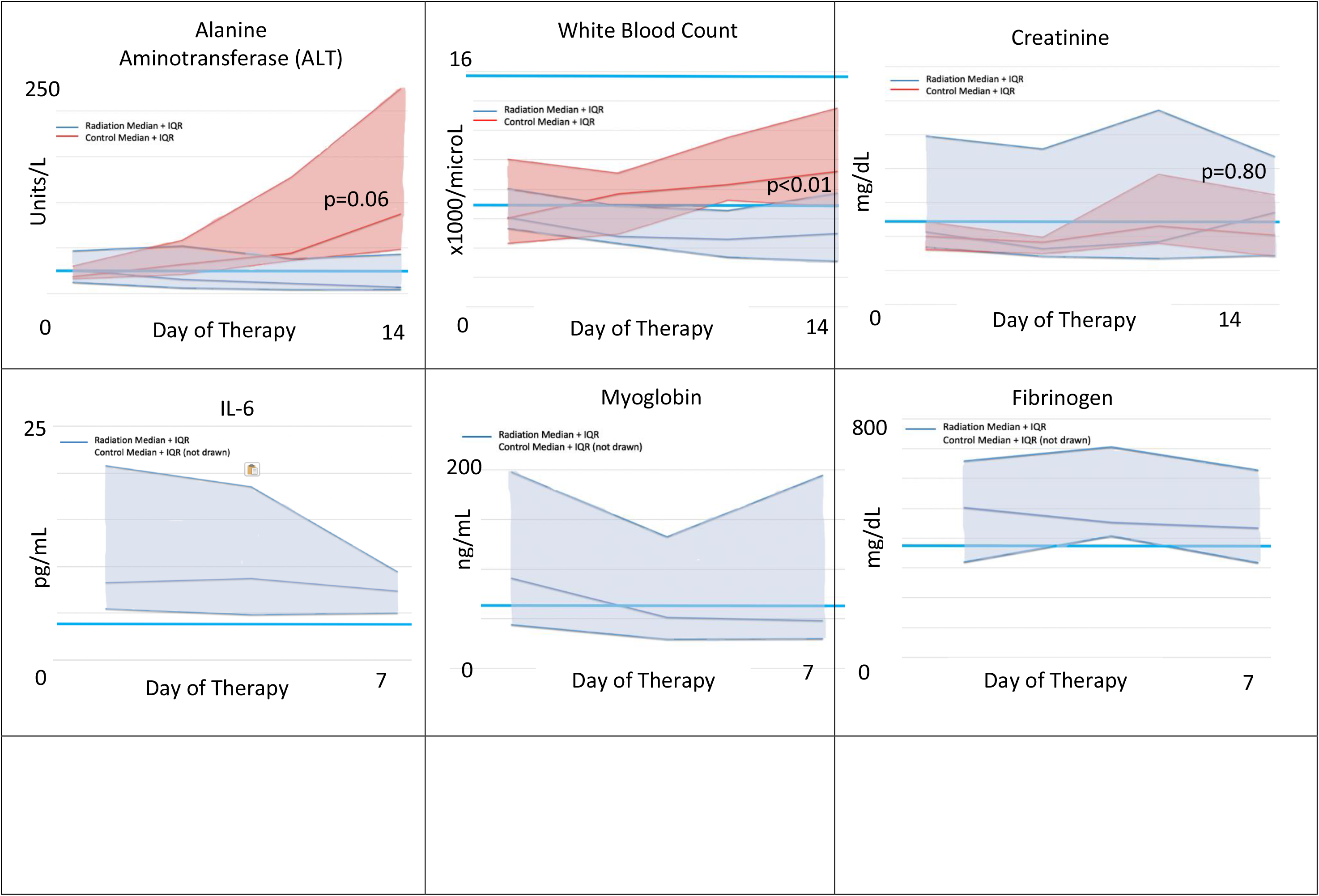
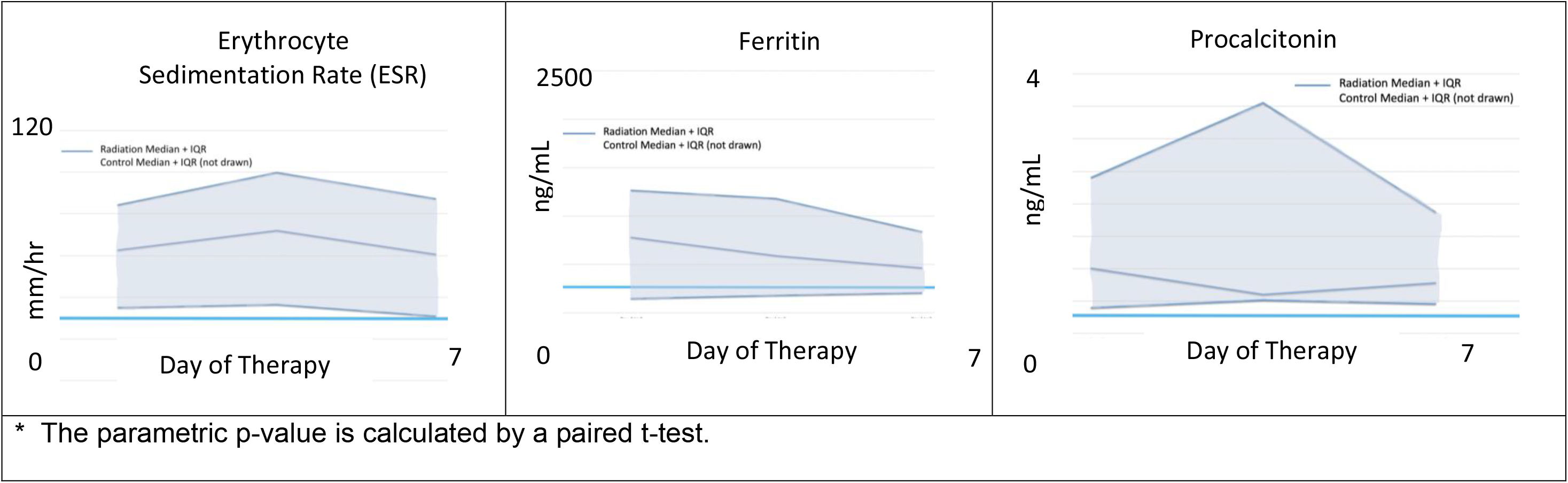
Serologic Marker Median and Interquartile Range Following Low-Dose Radiation Therapy Compared at Day 7 to Age- and Comorbidity-Matched Controls

### Adverse Events

One patient (10%) experienced CTCAE grade 1 upper gastrointestinal acute toxicity within 24 hours follow LD-RT delivery (nausea without alteration in eating habits). Another patient (patient 5) who presented with rapidly-increasing oxygenation requirements (up to 6L), required high flow oxygen support for 4 days following LD-RT. This was followed by systemic coagulation, cardiac, and renal lab abnormalities, intubation 5 days following LD-RT, and death on post-LD-RT day 15. Day 28 overall survival was 90% with 1 death in the LD-RT group. No other toxicity, airway emergencies, or other adverse events were observed following LD-RT.

## Discussion

This report describes 28-day outcomes of the phase II portion of the first reported trial exploring the efficacy of single-fraction, low-dose, whole-lung radiation for patients with COVID-19 pneumonia. In a cohort of only 10 patients, the effect of LD-RT was sufficiently large to be associated with a shorter time to clinical recovery of 3 versus 12 days in a cohort of age- and comorbidity-matched controls [HR 2.9, p=0.05]. Median time from intervention to hospital discharge was shorter at 12 days with LD-RT versus 20 days in the control group, and total hospital duration was shorter at 16 versus 19 days. Intubation was lower at 10% versus 40%, and total ICU stay was shorter at 10 versus 43 days. LD-RT yielded radiographic improvement and serologic recovery even in patients with high COVID burden at around 7 days, even after blinding the radiologist (BW) to the clinical intervention and timing thereof. Randomized evaluation of LD-RT is warranted to confirm these provocative results. Similar to dexamethasone, LD-RT may improve outcomes by reducing the virally-induced, hyperinflammatory response. Anti-Inflammatory effects of LD-RT can result in apoptosis and decrease adhesion of the leukocytes to endothelial cells,^10^ mitigate proinflammatory effects of macrophages by reducing secretion of nitric oxide and reactivation oxygen species,^11^ and induce polarization of M1-inflammatory macrophages to M2-anti-inflammatory subtype.^12^ LD-RT may also reduce TNF-α, IL-1β, IL-2, IL-6, IL-8, and INF-γ,^13-17^ which are thought to be increased in COVID-19 cytokine storm. While these findings require further investigation, whole-lung LD-RT did not induce post-treatment pancytopenia or immunosuppression and therefore is unlikely to slow viral clearance. In contrast, global immunosuppression, which dexamethasone may induce, slows viral clearance in murine models and remains a concern despite improving survival.^21,22^ Thus, it is plausible that radiotherapy may be additive to the effect of steroids in pre-intubated, hospitalized, and oxygen-dependent COVID-19 patient, who showed the least benefit following steroid administration, with only 1 in 25 deaths being prevented with steroids.^23,24^ The active phase 3 trial will compare LD-RT versus physicians’ choice of COVID-directed therapies in these patients.

### Potential Impact

As of June 2020, more than 8.9 million people globally are confirmed as infected with COVID-19, leading to over 466,000 known deaths. This report suggests the potential ability to improve upon the results of recent randomized trials with a 10-minute treatment that carries minimal toxicity and is well tolerated even in the elderly and fragile patients. Limitations to this study includes the non-randomized approach, small patient numbers, non-contemporaneous controls, limited imaging and serological studies in the control cohort beyond 7 days, and lack of detailed viral load evaluations in the LD-RT and control cohorts. Future work with LD-RT will include detailed CD-8 T-cell activation studies, CD-4 T cell activation, changes in B-cell profiles, antibody formation, and neutralization tests. This further immunological analysis will provide additional insights regarding the role of LD-RT to not only improve clinical outcomes, but perhaps aid viral clearance.

## Conclusion

A predominantly elderly hospitalized COVID-19 pneumonia patient cohort with oxygen dependence, visible pneumonic infiltrates, and clinical decline were recovered to room air at a median time of 3 days and discharged at a median time of 12 days, with rapid improvement in altered mental status by hour 24 and in radiographs by day 7 to 21. There was no significant acute toxicity, and comparison for efficacy against age- and comorbidity-matched controls showed a 3-fold improvement in time to clinical recovery. Ongoing international efforts to evaluate the optimal role of LD-RT in COVID-19 pneumonia are justified. Randomized evaluation in our phase 3 clinical trial is merited [Clinical Trial Registration: NCT04433949].

## Data Availability

All relevant data and results are included in the manuscript.

## Acknowledgements

We would like to acknowledge our collaborating physicians Dr. Jesse James MD, Dr. Michael Sterling MD, Dr. Charles Grodzin MD, Dr. Craig Coopersmith MD, Dr. Greg Martin MD, Dr. Marybeth Sexton MD MSC, Dr. Ramzy Rimawi MD, and Dr. Samer Melhem MD PhD for contributions and collaboration for trial accrual. Dr Srilatha Edupuganti MD, MPH for allowing access to the emergent phlebotomy protocol, Andrew Cheng for data management, Christopher Huerta – lab processing, Laurel Bristow, Kieffer Hellmeister, Nina McNair and Laila Hussaini for sample collection, Ellie Buttler-regulatory support.

